# Exploring participation bias in a home-based exercise and physical activity intervention study among kidney transplant recipients – an observational comparative study

**DOI:** 10.1101/2025.06.17.25329819

**Authors:** Jasmine De Beir, Marieke Vandecruys, Marie Renier, Veronique Cornelissen, Stefan De Smet, Sabina De Geest, Amaryllis H. Van Craenenbroeck, Diethard Monbaliu, Wim Van Biesen, Evi V. Nagler, Francis Verbeke, Karsten Vanden Wyngaert, Patrick Calders

**Author notes:** Corresponding author: (PC).

## Abstract

**Background:** Limited knowledge exists about individuals who decline participation in exercise trials. This study compared characteristics of participants and nonparticipants in a kidney transplant exercise trial using a dual-consent design.

**Methods:** The study was nested within the PHOENIX-Kidney randomized controlled trial, which evaluated a home-based exercise and physical activity intervention in kidney transplant recipients. At three months post-transplant, eligible individuals were invited to participate via two parallel consent pathways: one for trial participation and one for observational data of nonparticipants. Both groups completed self-reported questionnaires on sociodemographics, physical activity levels, and perceived motivators and barriers to physical activity. Reasons for nonparticipation were documented, and clinical data were extracted from medical records. Group comparisons used t-tests, Mann-Whitney U, chi-square, and Fisher’s exact tests.

**Results:** Of 345 screened kidney transplant recipients, 58% were eligible. Ineligibility was primarily due to medical contraindications (26%), language barriers (24%), and multi-organ transplantation (20%). Among eligible individuals, 88 (44%) enrolled in the intervention and 65 (35%) completed the observational follow-up for nonparticipants. Major reasons for nonparticipation included lack of time (23%), and feeling sufficiently active (20%). Nonparticipants reported higher physical activity levels (195 [120–360] vs. 103 [19–180] minutes per week; *P* = 0.003), while participants had higher motivator scores for physical activity (2.1 [1.6 – 2.4] vs. 1.5 [1.1 – 1.9]; *P* < 0.01).

**Conclusion:** The dual-consent approach enabled unique insights into differences between participants and nonparticipants, which can inform strategies to reduce participation bias and enhance inclusivity in future trials.

**Clinical Trial Registration:** Clinicaltrials.gov identifier number: NCT06260579.

## Introduction

Physical exercise training improves cardiopulmonary health, overall physical function, and quality of life in kidney transplant recipients (KTRs) [1]. However, low inclusion rates in randomized controlled trials (RCTs), the gold standard for evaluating intervention efficacy and effectiveness, raise concerns about external validity. Previous studies report that up to half of eligible KTRs decline participation in supervised exercise trials, citing travel distance, logistical challenges, lack of interest, time constraints, and occupational demands [1–4]. These factors highlight recruitment challenges and points to concerns regarding generalizability. Yet, the characteristics of nonparticipants remain underexplored.

Research in other clinical populations, such as individuals with cancer and rheumatoid arthritis, suggests that nonparticipants in exercise studies often face socioeconomic, disease-related, and psychological barriers. Compared to participants, they tend to report lower physical activity levels, poorer health-related quality of life, greater perceived disease burden, and more negative attitudes toward exercise [5, 6]. These findings emphasize the complexity of decision-making when patients consider engaging in physical activity interventions.

Among KTRs, observational studies have identified various barriers and motivators influencing engagement in physical activity and exercise [7]. Commonly reported barriers include concerns about harming the graft, feeling too unwell to exercise, lack of motivation or guidance, bad weather, fatigue, and accessibility issues [8–11]. Conversely, motivators such as enjoyment, health benefits, social interaction, support from healthcare professionals, and goals-setting have also been reported [9]. Transplant recipients who do not partake in exercise interventions generally report lower activity levels and higher perceived barriers compared to participants [11]. Together, these findings underscore the need to better understand the differences between those who participate in exercise trials and those who do not.

Direct comparisons between RCT participants and nonparticipants are typically limited by ethical restrictions on data collection from individuals who decline study participation. Methodological innovation is therefore needed to address this gap. A parallel study design that incorporates informed consent from both participants and nonparticipants offers a potential solution to enable such comparative analysis.

The aim of this study is to identify and compare characteristics of participants and nonparticipants in a kidney transplant exercise RCT, using a novel dual informed consent approach to enable direct comparison.

## Materials and methods

### Study design and participants

This observational comparative study is nested in the ongoing PHOENIX-Kidney study, which is a multicenter RCT evaluating the impact of a home-based exercise and physical activity intervention in KTRs (ClincalTrials.gov ID: NCT06260579 [12]) (Fig 1) [13]. Ethical approval for the PHOENIX-Kidney RCT was obtained from the local ethics committees of Leuven and Ghent University Hospitals (B3222022000937), and the study complies with the International Conference for Harmonization of Good Clinical Practice guidelines and the declaration of Helsinki.

**Fig 1.**
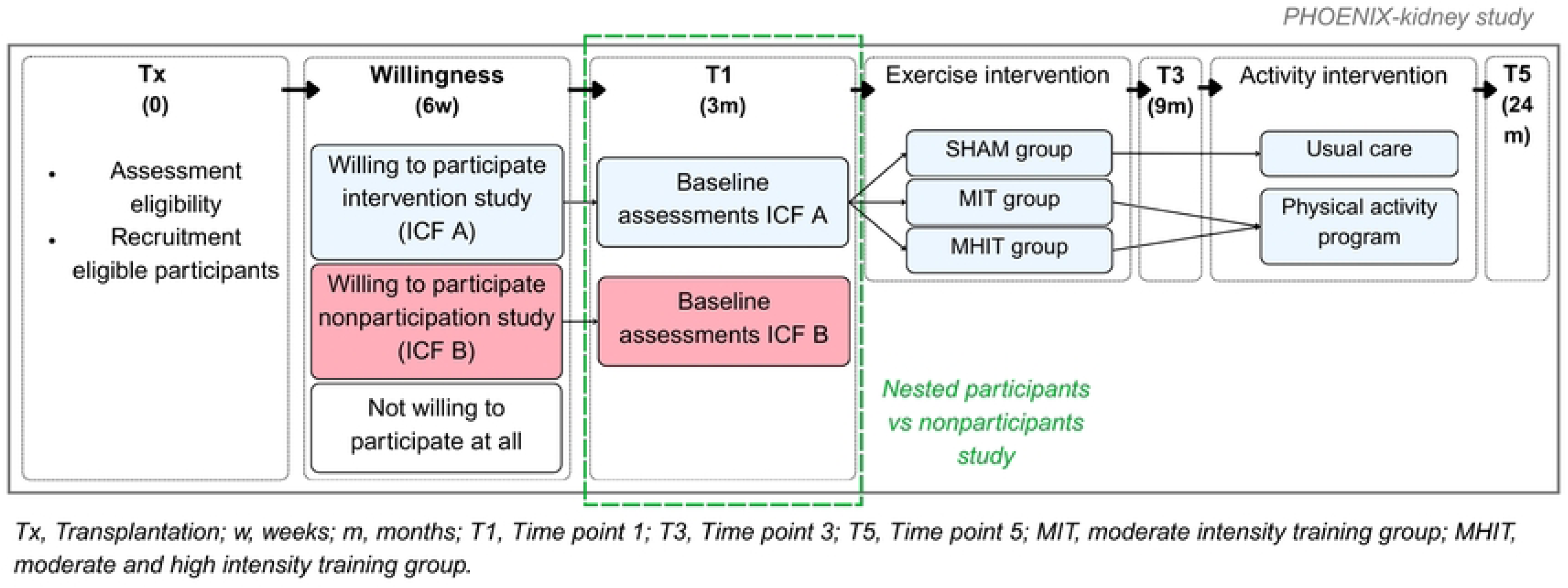
Nested study design of the participants vs nonparticipants study.

In brief, all de novo adult KTRs (aged ≥ 18 years) who underwent kidney transplantation at University Hospitals Leuven and Ghent in Belgium were screened for eligibility for the PHOENIX-Kidney RCT using predefined inclusion and exclusion criteria (Table 1). During their initial hospital admission, a nephrologist informed all eligible patients about the study. Six weeks post-transplantation, patients were asked about their willingness to participate in the trial. Those who agreed to participate in the PHOENIX-Kidney trial provided written informed consent (ICF-A) and underwent medical screening, including a maximal cardiopulmonary exercise test (CPET) supervised by a cardiologist, before being randomised into one of three exercise intervention groups: (1) six months of low-intensity training (SHAM), (2) six months of moderate-intensity training (MIT), or (3) three months of moderate-intensity training followed by three months of high-intensity training (MHIT). After the six-month training period, participants in group two and three transitioned to a maintenance physical activity program during 15 months, co-developed by a trained researcher using motivational interviewing and behavior change techniques. Participants in the SHAM group did not receive further physical activity guidance after completing their program.

**Table 1.**
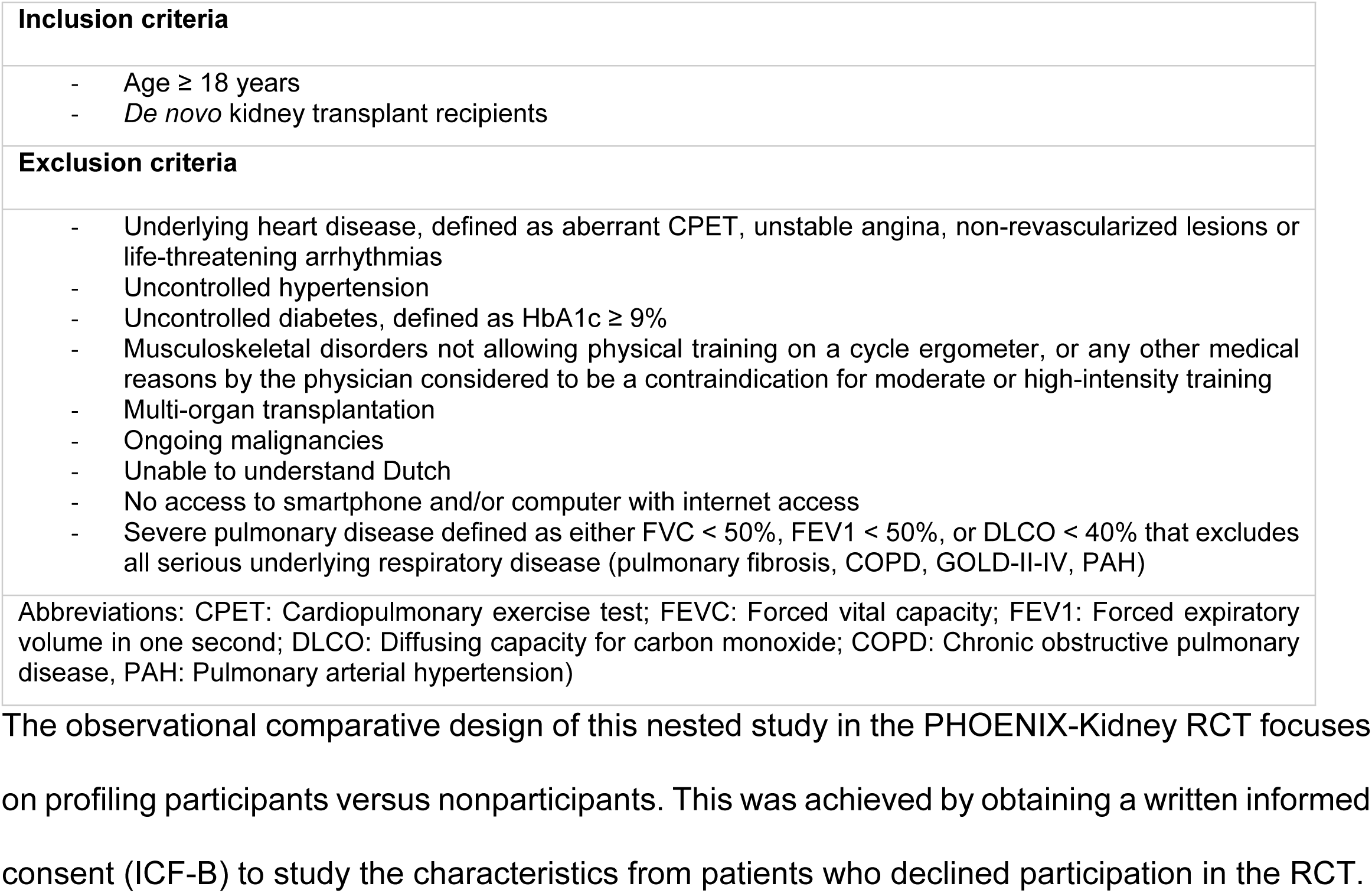
In– and exclusion criteria.

|  |
| --- |
| **Inclusion criteria** |
| - Age ≥ 18 years<br>- *De novo* kidney transplant recipients |
| **Exclusion criteria** |
| - Underlying heart disease, defined as aberrant CPET, unstable angina, non-revascularized lesions or life-threatening arrhythmias<br>- Uncontrolled hypertension<br>- Uncontrolled diabetes, defined as HbA1c ≥ 9%<br>- Musculoskeletal disorders not allowing physical training on a cycle ergometer, or any other medical reasons by the physician considered to be a contraindication for moderate or high-intensity training<br>- Multi-organ transplantation<br>- Ongoing malignancies<br>- Unable to understand Dutch<br>- No access to smartphone and/or computer with internet access<br>- Severe pulmonary disease defined as either FVC < 50%, FEV1 < 50%, or DLCO < 40% that excludes all serious underlying respiratory disease (pulmonary fibrosis, COPD, GOLD-II-IV, PAH) |
| Abbreviations: CPET: Cardiopulmonary exercise test; FEVC: Forced vital capacity; FEV1: Forced expiratory volume in one second; DLCO: Diffusing capacity for carbon monoxide; COPD: Chronic obstructive pulmonary disease, PAH: Pulmonary arterial hypertension) |

### Variables and measurement

Following variables were collected at three months post transplantation in both groups (ICF-A and ICF-B). Variables were selected to pragmatically explore factors associated with participation in exercise interventions, informed by previous literature [11].

#### Sociodemographic and Clinical characteristics

Sociodemographic data assessed through structured self-report questionnaire were sex, age, ethnicity, religion, living situation, informal care situation, family status, caregiving responsibilities, education, work status, income, savings, smoking habits, and alcohol consumption (questionnaire A in S1 Protocol). Clinical data extracted from the hospital’s electronic health records were following: donor type, cause of renal failure, pre-transplant dialysis vintage, estimated glomerular filtration rate (eGFR), height, weight and comorbidities. The comorbidity profile was assessed using the Functional Comorbidity Index (FCI) [14], which includes 18 medical conditions associated with physical functioning. Each condition was scored as present (1) or absent (0), yielding a total score ranging from 0 to 18.

#### Physical Activity Levels

Physical activity levels were self-reported using the Physical Activity Vital Sign (PAVS) questionnaire, which measured frequency and duration of moderate and vigorous physical activity, as well as the number of days per week engaged in strength training exercises [15]. Moderate and vigorous physical activity duration was reported in minutes per week, while strength training was reported in days per week. Based on the World Health Organization (WHO) guidelines, minimum 150 minutes of moderate to vigorous physical activity and minimum two days of strength training per week were used as cutoff values [16]. The PAVS questionnaire has previously demonstrated a moderate correlation (r = 0.52, *P* < .001) to accelerometer data, and a moderate agreement (κ = 0.46, *P* < .001) for categorizing participants’ physical activity levels based on meeting the recommended guidelines [17].

#### Barriers and Motivators to Physical Activity

Barriers and motivators to physical activity were measured using the self-reported Barriers and Motivators Questionnaire, a validated self-reported instrument that quantifies 31 barriers and 23 motivators on a 4-point scale (0 = ‘not at all’, to 3 = ‘very much’). This questionnaire, initially developed for hemodialysis patients [18], has also been used in studies involving solid organ transplant recipients [10, 11, 19, 20]. The barrier scale demonstrated good internal consistency (Cronbach’s α = 0.87), while the motivator scale showed excellent internal consistency (Cronbach’s α = 0.91). Given these high reliability values, arithmetic mean scores were calculated for both scales by averaging responses across all items. Additionally, percentages of positive answers on individual items were calculated based on responses of ‘slightly’ or higher.

#### Reasons for nonparticipation

Self-reported reasons for nonparticipation in the PHOENIX-Kidney trial were assessed with eight predefined options: *‘Lack of motivation,’ ‘Lack of transportation,’ ‘Lack of time,’ ‘Afraid the exercise program will be too strenuous,’ ‘Fear of injury,’ ‘Fatigue,’ ‘I do not like exercising/sports,’* and *‘I prefer not to give a reason’*. Participants were instructed to select only one primary reason, with an open-ended option to specify a reason not listed among the choices (questionnaire B in S1 Protocol). These options were self-composed based on common barriers identified in the literature [10, 21].

## Data collection

Data collection for the nested study took place between 30 November 2022 and 16 October 2024. Data were collected at baseline (T1), three months post-transplantation, for both participants and nonparticipants. Questionnaires were self-administered digitally or on paper, at home or in the hospital, based on participant preference. Participants could contact the research team with any questions. Additional study data were collected and managed in secure web-based platform (REDCap), ensuring secure and efficient data capture [22, 23].

## Statistical analyses

Descriptive statistics used for continuous variables were mean ± standard deviation (SD) or, median with interquartile ranges (IQR), depending on distribution. Categorical variables were expressed as frequencies and percentages. Group comparisons for continuous variables used independent Student’s t-tests or Mann-Whitney U-tests, and categorical variables were analyzed using Chi-square or Fisher’s exact tests. Statistical significance was set at *P* < 0.05 (two-tailed).

To compare groups across the 31 barrier and 23 motivator items of the Motivators and Barriers Questionnaire (MBQ), Chi-square or Fisher’s exact tests were applied. The Benjamini-Hochberg procedure controlled for Type I error using a False Discovery Rate correction with an initial α of 0.05.

Missing data were assessed and reported. Complete case analysis was used, excluding cases with missing values on variables of interest. All analyses were conducted using IBM SPSS Statistics (Version 29).

## Results

### Participant flow and recruitment

A total of 345 KTRs were screened for eligibility. Of these, 200 (58%) were eligible for study participation, while 145 (42%) were excluded due to: medical reasons contraindicating exercise training (25.5%), inability to understand Dutch (24%), multi organ transplantation (20%), post-transplant complications (16.5%), musculoskeletal disorders (9%), or organizational barriers (5%) (Fig 2).

**Fig 2.**
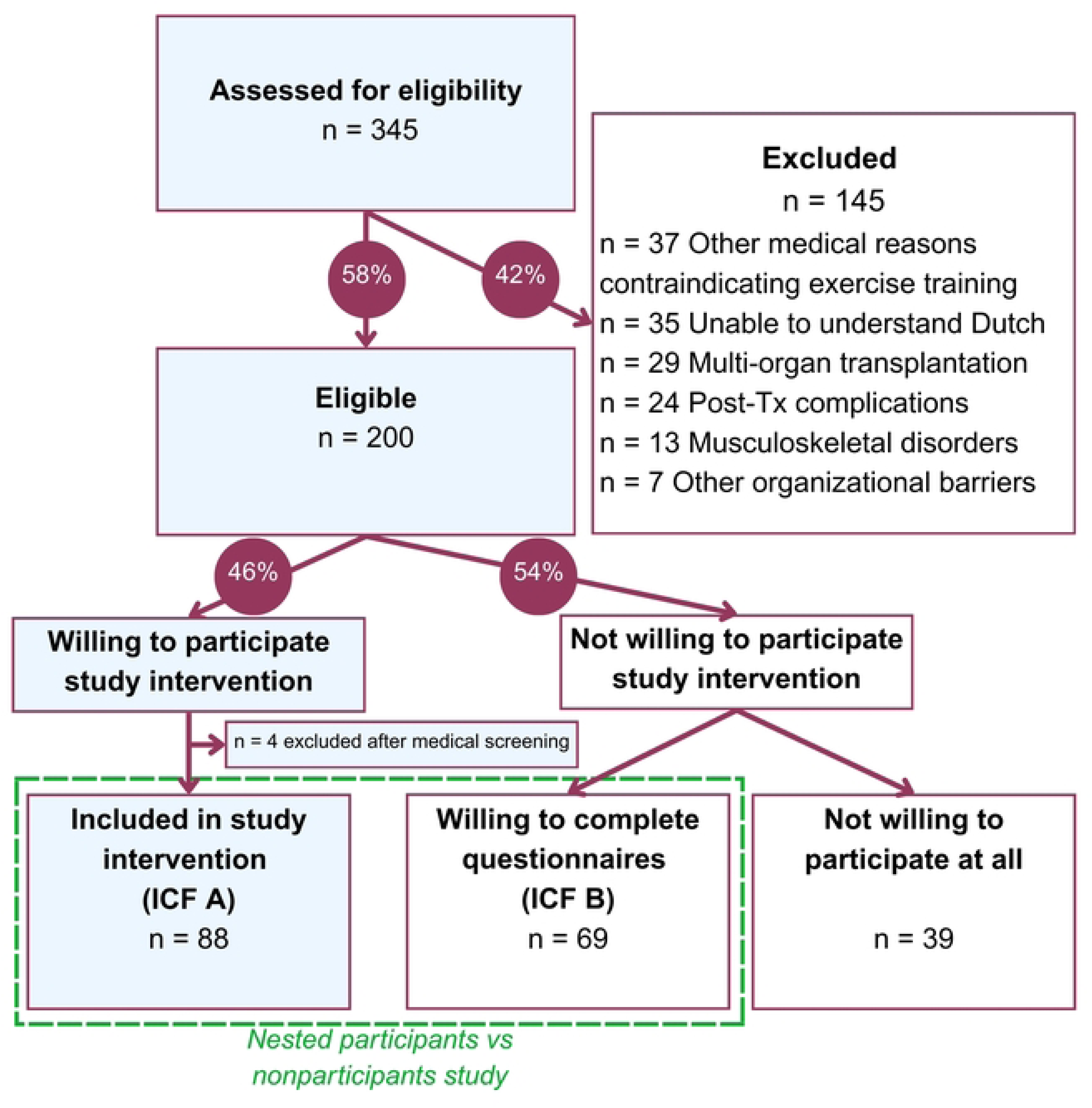
PHOENIX-Kidney flowchart.

Among eligible participants, 92 (46%) consented to participate in the PHOENIX-Kidney trial. Four participants were excluded after medical screening, resulting in a total of 88 individuals being randomized into one of three study arms (ICF-A). Among the 108 individuals who declined participation in the RCT, 69 (64%) agreed to partake in the nonparticipation study (ICF-B).

### Sociodemographic and clinical characteristics

Sociodemographic and clinical characteristics of participants and nonparticipants are summarized in Table 2. Overall, female individuals appeared to be less likely to participate in the RCT compared to male individuals, although this difference was not statistically significant (*P* = 0.07). No other meaningful differences were observed between the two groups.

**Table 2.** Sociodemographic and clinical characteristics of participants (ICF-A) and nonparticipants (ICF-B).

| Variable |  | Participants<br>ICF-A (n=88) | Nonparticipant<br>s ICF-B (n=69) | P-value |
| --- | --- | --- | --- | --- |
| **Transplantation center**<br>**(n, %)** | Leuven | 59 (67.0%) | 42 (60.9%) | 0.42 |
|  | Ghent | 29 (33.0%) | 27 (39.1%) |  |
| **Sex**<br>**(n, %)** | Male | 61 (69.3%) | 38 (55.1%) | 0.07 |
|  | Female | 27 (30.7%) | 31 (44.9%) |  |
| **Age (median, [IQR])** |  | 57.5 [47.3 - 64.0] | 57.0 [50.5 - 64.5] | 0.50 |
| **Cause of renal failure**<br>**(n, %)** | Glomerular disease | 20 (22.7%) | 24 (34.8%) | 0.25 |
|  | Familial/hereditary nephropathies | 30 (34.1%) | 20 (29.0%) |  |
|  | Other causes | 38 (43.2%) | 25 (36.2%) |  |
| **Pretransplant dialysis**<br>**(n, %)** | No | 9 (10.2%) | 6 (8.7%) | 0.77 |
|  | Yes | 46 (89.8%) | 62 (89.9%) |  |
|  | Missing |  | 1 (1.4%) |  |
| **Dialysis vintage in weeks (median [IQR])** |  | 127 [67 - 220] | 149 [66 - 241] | 0.44 |
| **Organ source**<br>**(n, %)** | Deceased | 73 (83.0%) | 61 (88.4%) | 0.34 |
|  | Living | 15 (17.0%) | 8 (11.6%) |  |
| **BMI [kg/m<sup>2</sup>] (mean ± SD)** |  | 24.7 ± 4.7 | 24.7 ± 4.4 | 0.93 |
| Missing (n) |  | 6 | 5 |  |
| **Functional Comorbidity Index (median, [IQR])** |  | 1 [0 - 2] | 1 [0 - 2] | 0.47 |
| **eGFR (ml/min/1.73m<sup>2</sup>) (mean ± SD)** |  | 49.4 [37.0 - 64.8] | 50.0 [33.0 - 60.2] | 0.58 |
| **Ethnic background**<br>**(n, %)** | White/East and West European | 77 (87.5%) | 68 (98.6%) | 1 |
|  | Other | 3 (3.3%) | 1 (1.4%) |  |
|  | Missing | 8 (9.1%) |  |  |
| **Religion**<br>**(n, %)** | Atheism | 25 (28.4%) | 20 (29.0%) | 0.83 |
|  | Christian | 48 (54.5%) | 45 (65.2%) |  |
|  | Other | 6 (6.8%) | 3 (4.3%) |  |
|  | Missing | 9 (10.2%) | 1 (1.4%) |  |
| **Living situation**<br>**(n, %)** | Alone | 17 (19.3%) | 14 (20.3%) | 1 |
|  | With others | 62 (70.5%) | 55 (79.7%) |  |
|  | Other | 1 (1.1%) |  |  |
|  | Missing | 8 (9.1%) |  |  |
| **Informal care situation**<br>**(n, %)** | No | 6 (6.8%) | 2 (2.9%) | 0.29 |
|  | Yes | 74 (84.1%) | 67 (97.1%) |  |
|  | Missing | 8 (9.1%) |  |  |
| **Family status**<br>**(n, %)** | No children | 17 (19.3%) | 20 (29%) | 0.53 |
|  | 1 child | 16 (18.2%) | 11 (15.9%) |  |
|  | 2 children | 28 (31.8%) | 27 (39.1%) |  |
|  | ≥ 3 children | 18 (20.5%) | 11 (15.9%) |  |
|  | Missing | 9 (10.2%) |  |  |
| **Caregiving**<br>**responsibilities (n, %)** | No | 43 (48.9%) | 33 (47.8%) | 0.47 |
|  | Yes | 37 (42.0%) | 36 (52.2%) |  |
|  | Missing | 8 (9.1%) |  |  |
| **Education**<br>**(n, %)** | None or primary | 5 (5.7%) | 8 (11.6%) | 0.19 |
|  | Secondary | 47 (53.4%) | 46 (66.7%) |  |
|  | Tertiary (Higher education) | 26 (29.5%) | 15 (21.7%) |  |
|  | Other | 2 (2.3%) |  |  |
|  | Missing | 8 (9.1%) |  |  |
| **Occupation**<br>**(n, %)** | Self-employed | 7 (8.0%) | 2 (2.9%) | 0.60 |
|  | Employed | 7 (8.0%) | 7 (10.1%) |  |
|  | House(wo)man | 4 (4.5%) | 8 (11.6%) |  |
|  | Retired | 18 (20.5%) | 14 (20.3%) |  |
|  | Sick/disability leave | 38 (43.2%) | 31 (44.6%) |  |
|  | Other | 4 (4.5%) | 2 (2.9%) |  |
|  | Missing | 5 (5.7%) | 5 (7.2%) |  |
| **Income**<br>**(n, %)** | <2000 euro/month | 46 (52.3%) | 37 (53.6%) | 0.88 |
|  | 2000-3000 euro/month | 24 (27.3%) | 22 (31.9%) |  |
|  | >3000 euro/month | 6 (6.8%) | 4 (5.8%) |  |
|  | Missing | 12 (13.6%) | 6 (8.7%) |  |
| **Savings**<br>**(n, %)** | <400 euro/month | 63 (71.6%) | 53 (76.8%) | 0.82 |
|  | ≥400 euro/month | 14 (15.9%) | 13 (18.8%) |  |
|  | Missing | 11 (12.5%) | 3 (4.3%) |  |
| **Smoking habits**<br>**(n, %)** | I don't smoke | 66 (75%) | 56 (81.2%) | 0.83 |
|  | I stopped smoking | 14 (15.9%) | 13 (18.8%) |  |
|  | Missing | 8 (9.1%) |  |  |
| **Alcohol consumption**<br>**(n, %)** | ≤3 days/month | 63 (71.6%) | 60 (87.0%) | 0.15 |
|  | ≥1 day/week | 18 (20.5%) | 9 (13.0%) |  |
|  | Missing | 7 (8.0%) |  |  |

Abbreviations: n, number; SD, standard deviation; BMI, body mass index; eGFR, estimated glomerular filtration rate.

### Physical activity levels

Self-reported physical activity levels varied significantly between participants (ICF-A) and nonparticipants (ICF-B) (Table 3). Nonparticipants reported notably higher activity levels, with 67% meeting the WHO recommendations of at least 150 minutes of moderate-to-vigorous physical activity per week, compared to 35% of participants. However, strength training engagement was low in both groups, with only about a quarter of individuals in each group meeting the recommendations of at least two training sessions per week.

**Table 3.**
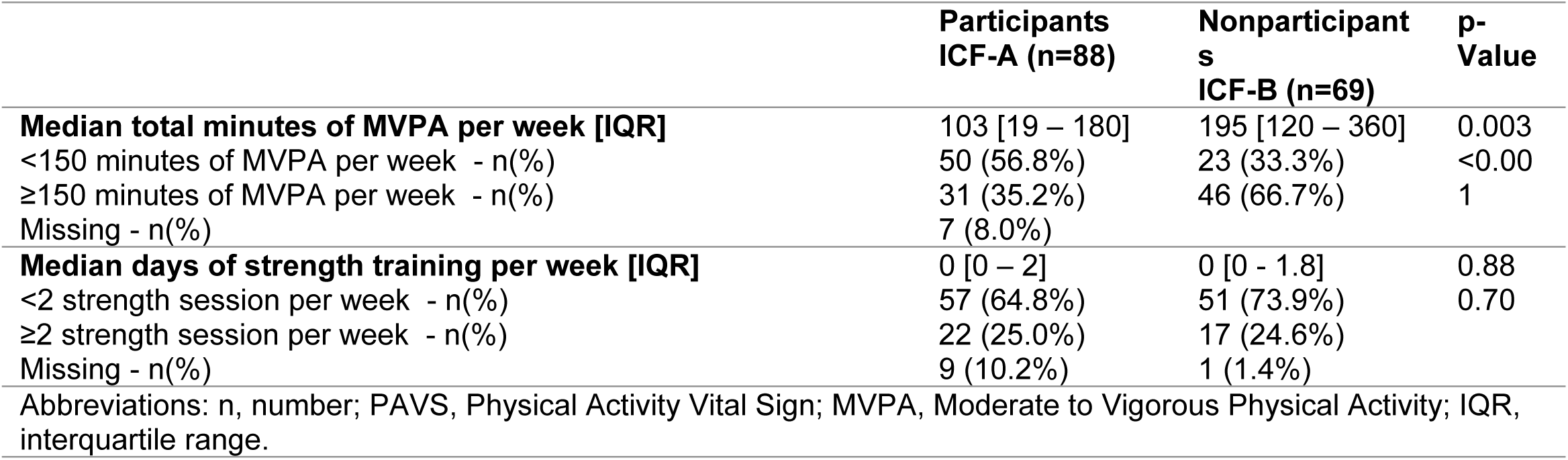
Physical activity levels of participants (ICF-A) and nonparticipants (ICF-B) according to the PAVS.

### Mean motivator and barrier scores

Mean motivator and barrier scores to physical activity (MBQ) are visualized in Fig 3. Participants (ICF-A) reported higher mean motivator scores compared to nonparticipants (ICF-B) (ICF-A: 2.1 [1.6–2.4] vs. ICF-B: 1.5 [1.1–1.9]), while mean barrier scores were low and similar between the two groups (ICF-A: 0.3 [0.2–0.6] vs. ICF-B: 0.3 [0.2–0.6]).

**Fig 3.**
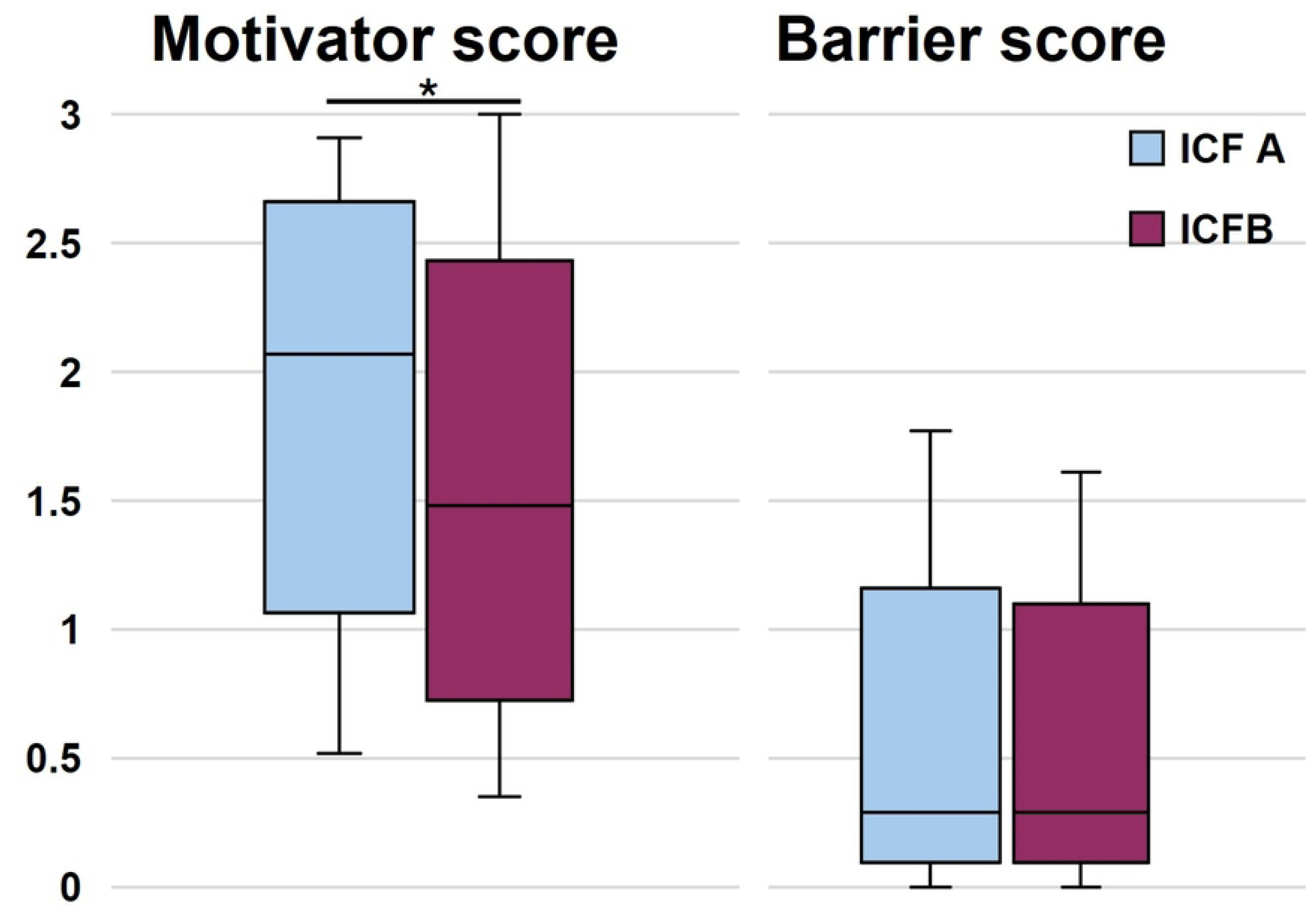
Motivator and Barrier scores to physical activity for participants (ICF-A) and nonparticipants (ICF-B) expressed as boxplots. \**P* < 0.05.

### Prevalence of individual motivator and barrier items (%)

Analysis of individual motivator statements for physical activity showed that a statistically significantly higher percentage of participants (ICF-A) identified certain factors as more motivating compared to nonparticipants (ICF-B) (Fig 4). Other motivator items showed no difference between both groups. Health benefits were the most commonly recognized motivators in both groups, with improved health cited by 100% of participants and 98.5% of nonparticipants (Table A in S2 Table).

**Fig 4.**
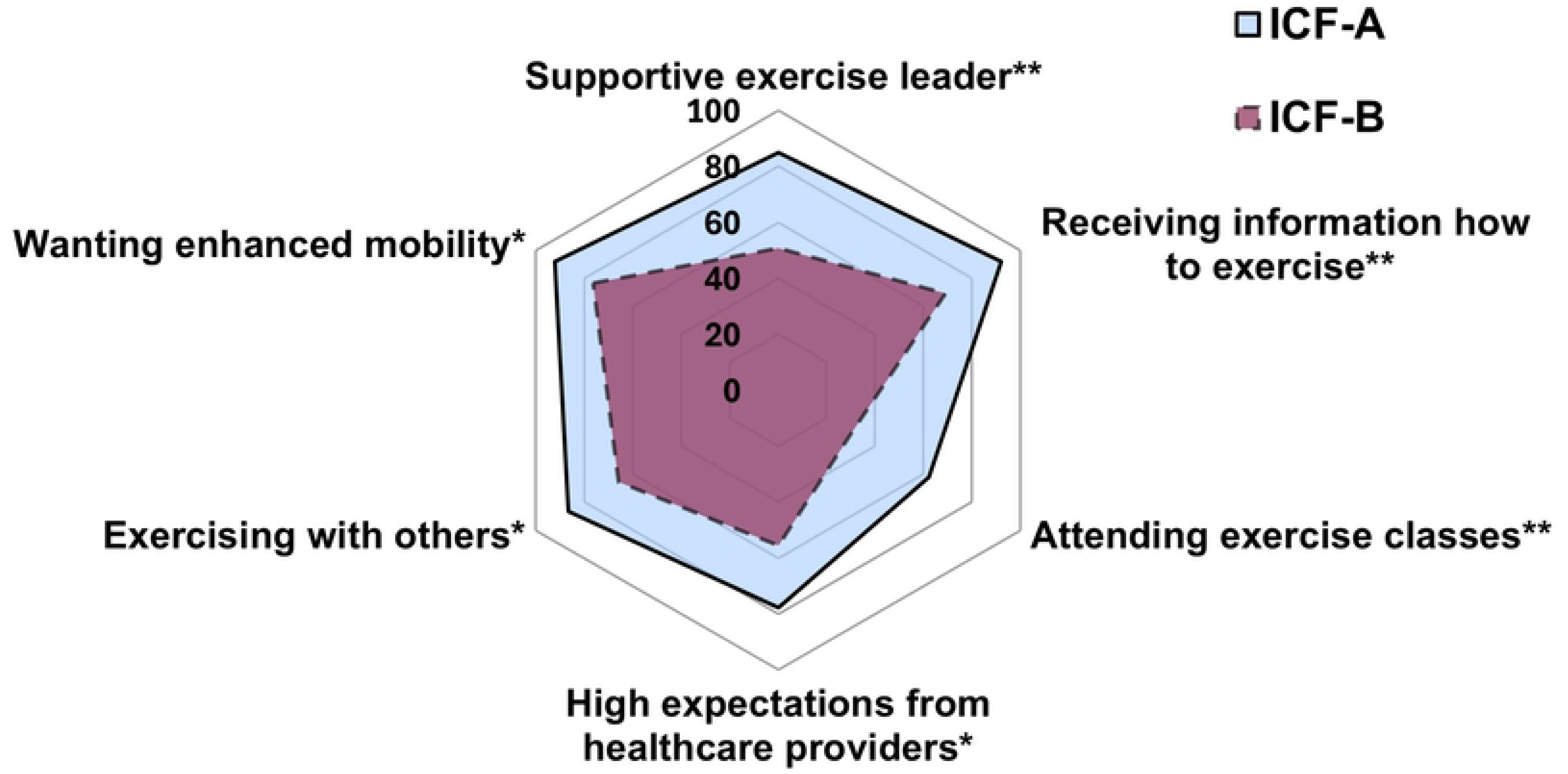
Hexagon visualising differences in motivators to physical activity between participants (ICF-A) and nonparticipants (ICF-B). \**P* < 0.05, \*\**P* < 0.01.

Perceived barriers did not differ between the two groups. Fatigue was the most frequently reported barrier, affecting 74 % of participants (ICF-A) and 63 % of nonparticipants (ICF-B) (Table B in S2 Table).

### Reasons for nonparticipation

Reasons for nonparticipation in the PHOENIX-Kidney trial are shown in Table 4. The most common reasons were lack of time and perceiving their physical activity levels as sufficient. Other reasons included concerns about the intervention format and various personal barriers.

**Table 4.**
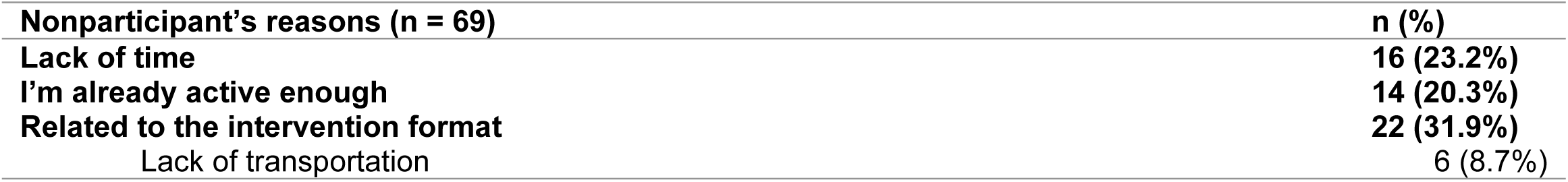
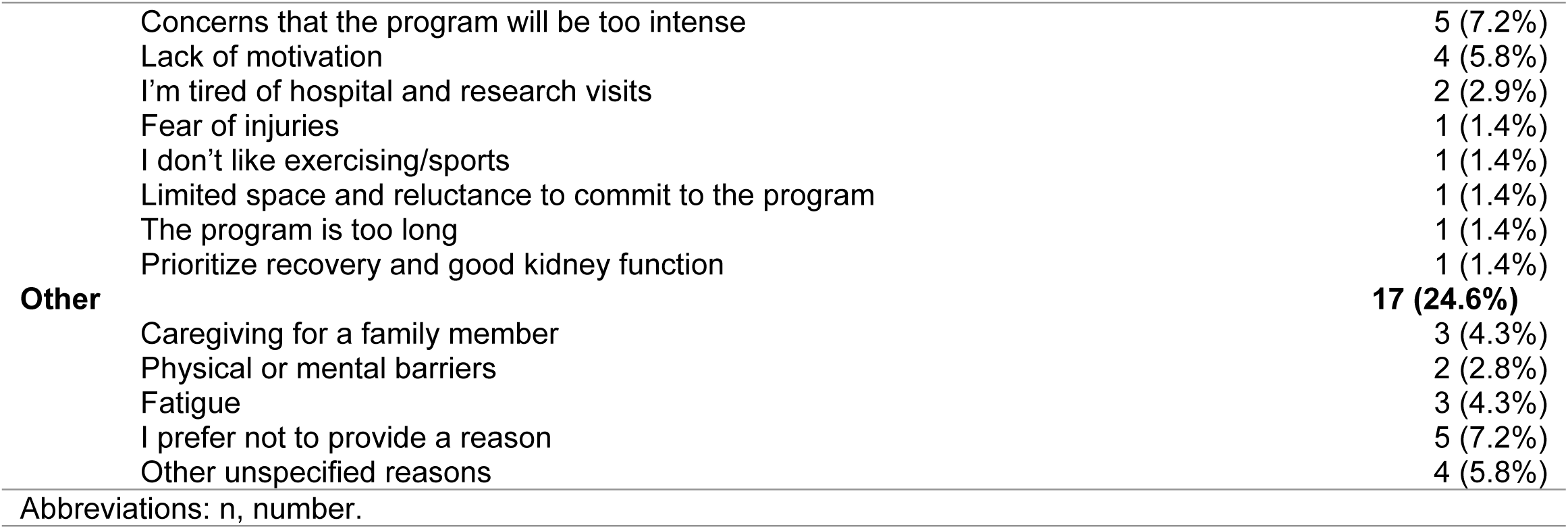
Reasons given by nonparticipants for declining to participate in the study.

| Nonparticipant's reasons (n = 69) | n (%) |
| --- | --- |
| Lack of time | 16 (23.2%) |
| I'm already active enough | 14 (20.3%) |
| Related to the intervention format | 22 (31.9%) |
| Lack of transportation | 6 (8.7%) |

|  |  |  |
| --- | --- | --- |
|  | Concerns that the program will be too intense | 5 (7.2%) |
|  | Lack of motivation | 4 (5.8%) |
|  | I'm tired of hospital and research visits | 2 (2.9%) |
|  | Fear of injuries | 1 (1.4%) |
|  | I don't like exercising/sports | 1 (1.4%) |
|  | Limited space and reluctance to commit to the program | 1 (1.4%) |
|  | The program is too long | 1 (1.4%) |
|  | Prioritize recovery and good kidney function | 1 (1.4%) |
| **Other** |  | **17 (24.6%)** |
|  | Caregiving for a family member | 3 (4.3%) |
|  | Physical or mental barriers | 2 (2.8%) |
|  | Fatigue | 3 (4.3%) |
|  | I prefer not to provide a reason | 5 (7.2%) |
|  | Other unspecified reasons | 4 (5.8%) |

Abbreviations: n, number.

## Discussion

This study is the first to characterize and compare participants and nonparticipants in a home-based exercise and physical activity intervention following kidney transplantation, using a parallel design with dual informed consent. Nonparticipants noted higher self-reported physical activity levels, whereas participants identified more motivators to engage in physical activity.

### Participation rate

In this study, 27% of the total approached population were both eligible and willing to participate in the exercise intervention, aligning with previous findings in KTRs [2–4, 24–31]. The main reasons for nonparticipation among eligible individuals included lack of time, perceived sufficient activity levels, logistical issues, and personal health concerns. Logistic issues and lack of time are well-documented barriers in earlier research [2–4]. In this study, a notable proportion of nonparticipants cited already being sufficiently active as a reason for nonparticipation, reflecting findings from a previous exercise and diet intervention, where 19% of nonparticipants reported the same reason [3]. Interestingly, health concerns specifically related to participation in an exercise study have not been reported among KTRs in previous studies [2–4]. These findings underscore the need to identify and address perceived barriers early in the recruitment process to enhance participation.

### Demographics of participants and nonparticipants

Despite no differences in sociodemographic characteristics between participants and nonparticipants, the sample does not fully capture female KTRs, 31% of participants is female vs. 45% of nonparticipants. Their underrepresentation aligns with well-documented barriers that disproportionally affect women’s engagement in physical activity and exercise interventions, including limited family support, intrapersonal challenges, and negative perceptions of exercise programs [21, 32, 33]. Addressing these gender-specific obstacles is essential to ensure equitable access and meaningful participation for female KTRs.

### Physical Activity Levels and Barriers for Participation

Notably, a high proportion of nonparticipants (ICF-B) (66%) reported meeting or exceeding the WHO-recommended physical activity levels of at least 150 minutes of moderate to vigorous physical activity per week, with a median of 210 minutes. This aligns with activity levels in a Dutch KTR cohort [19], suggesting that many nonparticipants had already established exercise habits, reducing the appeal of structured interventions. In contrast, only 35% of participants met the recommended levels, indicating that the study effectively engaged individuals who could benefit most, rather than solely attracting already active individuals as reported in earlier studies [34].

Beyond physical activity levels, other reasons for nonparticipation included concerns about program intensity, duration, potential injuries, and prioritizing recovery post-transplantation. Although the intervention was tailored to individual fitness levels using cardiorespiratory exercise testing and included co-creating physical activity goals, some may have perceived it as too demanding or misaligned with their current health needs. Fatigue also emerged as a common barrier across groups; however, while participants may have viewed exercise as a strategy to reduce fatigue, nonparticipants may have perceived it as an additional burden. Further in-depth interviews are needed to explore this distinction. These insights underscores the importance of framing exercise as a possible solution to fatigue, given its documented benefits in KTRs [24], and adopting an individualized recruitment approach to address specific concerns.

### Intervention format and participation preferences

Participants perceived higher motivators to exercise, consistent with prior findings in solid organ transplant recipients [11]. They valued external support, including structured exercise sessions, guidance from an exercise coach, information on how to exercise, and high expectations from healthcare providers. In contrast, nonparticipants preferred independent physical activity. This preference mismatch aligned with evidence that those favoring self-directed exercise are less likely to engage in structured programs [35]. Additionally, reluctance toward randomization may also have discouraged some from participating [3, 36]. These insights highlight the challenges of recruiting diverse profiles into RCTs and underscore the need for flexible, participant-centered approaches in clinical exercise interventions.

Despite being home-based, the intervention did not fully eliminate logistical barriers; time constraints remaining a major obstacle, even among individuals previously undergoing time-intensive dialysis [37]. This suggests the post-transplant adjustment phase plays a crucial role [38], as patients increasingly value social, professional, and personal responsibilities, making these structured programs harder to prioritize. A similar dynamic was observed in stroke rehabilitation, where individuals must adapt to new roles [39]. To promote sustained participation, programs should incorporate personalized coaching to help integrate exercise into daily routines. Additionally, timing may be crucial: rather than a fixed start point post-transplantation, rehabilitation programs might be more effective if introduced once individuals have settled into their new roles. Further research should explore optimal timing and delivery of rehabilitation programs to align with patients’ evolving needs and daily life patterns.

### Strengths, limitations and future directions

This study is the first to provide a detailed comparison of participants and nonparticipants in an exercise intervention study following kidney transplantation. A key strength is the parallel design, allowing inclusion of nonparticipants to identify distinguishing characteristics. However, variable selection was not theory-driven, potentially overlooking relevant determinants. A major limitation is the reliance on self-reported physical activity, which may be susceptible to recall and social desirability biases, particularly among nonparticipants who may have overestimated their activity to justify nonparticipation. Prior research showed that self-reported activity in KTRs often exceed levels measured by accelerometery [19], raising concerns about accuracy. Selective nonresponse may also have occurred, with less active individuals being less likely to complete the survey. Additionally, missing data reduced completeness. Future research should incorporate objective activity measures and qualitative methods to better understand participation barriers.

### Clinical implications

Caution is needed when translating these findings into clinical practice, as the PHOENIX-Kidney study was an RCT with specific inclusion and exclusion criteria, selecting vital KTRs for safety reasons related to the high-intensity exercise intervention. Importantly, this observational study provided insights into why eligible individuals declined participation. By characterizing nonparticipants, it highlighted a range of barriers—such as sex, physical activity level, and health concerns—that may be addressed through personalized recruitment strategies. Tailoring communication and support to match the concerns and preferences of potential participants could enhance inclusivity and engagement. The MBQ may be a valuable tool for identifying individual motivators to support this personalized approach. Notably, the intervention successfully engaged individuals with low physical activity levels but high motivation—those who may benefit most— highlighting the potential clinical values of such programs. Further research is needed to optimize recruitment strategies and intervention design for broader implementation.

## Conclusion

This study highlights the value of examining both participants and nonparticipants in exercise interventions following kidney transplantation. While the intervention effectively reached individuals with low physical activity levels and high motivation, a substantial proportion of eligible individuals declined participation due to perceived sufficient activity, health concerns, time constraints, and a preference for independent exercise. These findings underscore the need for flexible, individualized recruitment strategies that address diverse barriers and preferences.

## Data Availability

All relevant data are within the manuscript and its Supporting Information files.

## Acknowledgements

The authors would like to thank the participants for their valuable contribution to this study. We also extend our gratitude to the University Hospitals of Ghent and Leuven for their support in facilitating this research. Acknowledgment is given to the use of ChatGPT (version GPT-4, OpenAI, https://www.openai.com/) as language assistance tool. Finally, we appreciate the Ethical Committees of both institutions for overseeing the ethical aspects of this study.

## Supporting information files

**S1 Protocol. Sociodemographic questionnaire and reasons for nonparticipation.**

**S2 Table. Perceived motivators and barriers to physical activity and exercise endorsed by ICF A and ICF B, ranked by prevalence (%).**

**S3 Checklist. STROBE checklist.**

## References

[1] De Smet S, Van Craenenbroeck AH. Exercise training in patients after kidney transplantation. Clinical Kidney Journal 2021; 14: II15–II24.

[2] Riess KJ, Haykowsky M, Lawrance R, et al. Exercise training improves aerobic capacity, muscle strength, and quality of life in renal transplant recipients. *Applied Physiology*, Nutrition and Metabolism 2014; 39: 566–571.

[3] Knobbe TJ, Kremer D, Zelle DM, et al. Effect of an exercise intervention or combined exercise and diet intervention on health-related quality of life-physical functioning after kidney transplantation: the Active Care after Transplantation (ACT) multicentre randomised controlled trial. Lancet Healthy Longev 2024; 5: 100622.

[4] Hernández Sánchez S, Carrero JJ, Morales JS, et al. Effects of a resistance training program in kidney transplant recipients: A randomized controlled trial. Scand J Med Sci Sports 2021; 31: 473–479.

[5] De Jong Z, Munneke M, Jansen LM, et al. Differences between participants and nonparticipants in an exercise trial for adults with rheumatoid arthritis. Arthritis Care Res (Hoboken) 2004; 51: 593–600.

[6] Vd Wiel HJ, Stuiver MM, May AM, et al. Characteristics of participants and nonparticipants in a blended internet-based physical activity trial for breast and prostate cancer survivors: Cross-sectional study. JMIR Cancer; 7. Epub ahead of print 1 October 2021. DOI: 10.2196/25464.

[7] Leunis S, Vandecruys M, Cornelissen V, et al. Physical Activity Behaviour in Solid Organ Transplant Recipients: Proposal of Theory-Driven Physical Activity Interventions. Kidney and Dialysis 2022; 2: 298–329.

[8] Gordon EJ, Prohaska TR, Gallant M, et al. Self-care strategies and barriers among kidney transplant recipients: A qualitative study. Chronic Illn 2009; 5: 75–91.

[9] Billany RE, Smith AC, Stevinson C, et al. Perceived barriers and facilitators to exercise in kidney transplant recipients: A qualitative study. Health Expectations 2022; 25: 764–774.

[10] Sánchez Z V., Cashion AK, Cowan PA, et al. Perceived barriers and facilitators to physical activity in kindey transplant recipients. Progress in Transplantation 2007; 17: 324–331.

[11] Masschelein E, De Smet S, Denhaerynck K, et al. Patient-reported outcomes evaluation and assessment of facilitators and barriers to physical activity in the Transplantoux aerobic exercise intervention. PLoS One; 17. Epub ahead of print 1 October 2022. DOI: 10.1371/journal.pone.0273497.

[12] Van Craenenbroeck AH. Home-based Exercise and Physical Activity Intervention After Kidney Transplantation: Impact of Exercise Intensity (PHOENIX-Kidney). ClinicalTrial.gov, https://clinicaltrials.gov/study/NCT06260579 (2024, accessed 15 April 2025).

[13] De Smet S, Vandecruys M, De Beir J, et al. Home-based exercise and physical activity intervention after kidney transplantation – impact of exercise intensity [PHOENIX-Kidney]. Clin Kidney J. Epub ahead of print 25 April 2025. DOI: 10.1093/ckj/sfaf114.

[14] Groll DL, To T, Bombardier C, et al. The development of a comorbidity index with physical function as the outcome. J Clin Epidemiol 2005; 58: 595–602.

[15] Golightly YM, Allen KD, Ambrose KR, et al. Physical Activity as a Vital Sign: A Systematic Review. Prev Chronic Dis; 14. Epub ahead of print 2017. DOI: 10.5888/pcd14.170030.

[16] World Health Organization. WHO GUIDELINES ON PHYSICAL ACTIVITY AND SEDENTARY BEHAVIOUR AT A GLANCE. Geneva, 2020.

[17] Ball TJ, Joy EA, Goh TL, et al. Validity of two brief primary care physical activity questionnaires with accelerometry in clinic staff. Prim Health Care Res Dev 2015; 16: 100–108.

[18] Goodman ED, Ballou MB. Perceived barriers and motivators to exercise in hemodialysis patients. Nephrol Nurs J 2004; 31: 23–9.

[19] Van Adrichem EJ, Dekker R, Krijnen WP, et al. Physical Activity, Sedentary Time, and Associated Factors in Recipients of Solid-Organ Transplantation, https://academic.oup.com/ptj/article/98/8/646/4994805 (2018).

[20] van Adrichem EJ, Krijnen WP, Dekker R, et al. Multidimensional structure of a questionnaire to assess barriers to and motivators of physical activity in recipients of solid organ transplantation. Disabil Rehabil 2017; 39: 2330–2338.

[21] Resurrección DM, Motrico E, Rigabert A, et al. Barriers for Nonparticipation and Dropout of Women in Cardiac Rehabilitation Programs: A Systematic Review. Journal of Women’s Health 2017; 26: 849–859.

[22] Harris PA, Taylor R, Minor BL, et al. The REDCap consortium: Building an international community of software platform partners. Journal of Biomedical Informatics; 95. Epub ahead of print 1 July 2019. DOI: 10.1016/j.jbi.2019.103208.

[23] Harris PA, Taylor R, Thielke R, et al. Research electronic data capture (REDCap)-A metadata-driven methodology and workflow process for providing translational research informatics support. J Biomed Inform 2009; 42: 377–381.

[24] SenthilKumar T, Soundararajan P, Maiya AG, et al. Effects of graded exercise training on functional capactiy, muscle strength, and fatigue after renal transplantation. Saudi Journal of Kidney Diseases and Transplantation 2020; 31: 100–108.

[25] Lima PS, De Campos AS, De Faria Neto O, et al. Effects of Combined Resistance Plus Aerobic Training on Body Composition, Muscle Strength, Aerobic Capacity, and Renal Function in Kidney Transplantation Subjects, www.nsca.com (2019).

[26] O’Connor EM, Koufaki P, Mercer TH, et al. Long-term pulse wave velocity outcomes with aerobic and resistance training in kidney transplant recipients – A pilot randomised controlled trial. PLoS One; 12. Epub ahead of print 1 February 2017. DOI: 10.1371/journal.pone.0171063.

[27] Karelis AD, Hébert MJ, Rabasa-Lhoret R, et al. Impact of Resistance Training on Factors Involved in the Development of New-Onset Diabetes After Transplantation in Renal Transplant Recipients: An Open Randomized Pilot Study. Can J Diabetes 2016; 40: 382– 388.

[28] Kouidi E, Vergoulas G, Anifanti M, et al. A randomized controlled trial of exercise training on cardiovascular and autonomic function among renal transplant recipients. Nephrology Dialysis Transplantation 2013; 28: 1294–1305.

[29] Zhang P, Liu S, Zhu X, et al. The effects of a physical exercise program in Chinese kidney transplant recipients: a prospective randomised controlled trial. Clin Kidney J 2023; 16: 1316–1329.

[30] Onofre T, Fiore Junior JF, Amorim CF, et al. Impact of an early physiotherapy program after kidney transplant during hospital stay: a randomized controlled trial. J Bras Nefrol 2017; 39: 424–432.

[31] Maia TO, Paiva DN, Sobral Filho DC, et al. Does whole body vibration training improve heart rate variability in kidney transplants patients? A randomized clinical trial. J Bodyw Mov Ther 2020; 24: 50–56.

[32] Murtagh EM, Murphy MH, Murphy NM, et al. Prevalence and correlates of physical inactivity in community-dwelling older adults in Ireland. PLoS One; 10. Epub ahead of print 11 February 2015. DOI: 10.1371/journal.pone.0118293.

[33] Wilkinson TJ, Clarke AL, Nixon DGD, et al. Prevalence and correlates of physical activity across kidney disease stages: An observational multicentre study. Nephrology Dialysis Transplantation 2021; 36: 641–649.

[34] Barreto P de S, Ferrandez A-M, Saliba-Serre B. Are Older Adults Who Volunteer to Participate in an Exercise Study Fitter and Healthier Than Nonvolunteers? The Participation Bias of the Study Population. J Phys Act Health 2013; 10: 359–367.

[35] Bisschop CNS, Courneya KS, Velthuis MJ, et al. Control group design, contamination and drop-out in exercise oncology trials: A systematic review. PLoS One; 10. Epub ahead of print 27 March 2015. DOI: 10.1371/journal.pone.0120996.

[36] Courneya KS, Forbes CC, Trinh L, et al. Patient satisfaction with participation in a randomized exercise trial: Effects of randomization and a usual care posttrial exercise program. Clinical Trials 2013; 10: 959–966.

[37] Kramer A, Boenink R, Mercado Vergara CG, et al. Time trends in preemptive kidney transplantation in Europe: an ERA Registry study. Nephrology Dialysis Transplantation. Epub ahead of print 27 November 2024. DOI: 10.1093/ndt/gfae105.

[38] De Beir J, De Baets S, Vandecruys M, et al. Challenges in posttransplantation care for kidney transplant recipients: A qualitative study highlighting gaps in psychological, social and exercise support. J Ren Care. Epub ahead of print 1 December 2024. DOI: 10.1111/jorc.12507.

[39] Hole E, Stubbs B, Roskell C, et al. The patient’s experience of the psychosocial process that influences identity following stroke rehabilitation: A metaethnography. The Scientific World Journal; 2014. Epub ahead of print 2014. DOI: 10.1155/2014/349151.

